# Using human genetics to validate complement C3 as a drug target in periodontitis

**DOI:** 10.1101/2023.02.13.23285838

**Authors:** Zoheir Alayash, Sebastian-Edgar Baumeister, Birte Holtfreter, Thomas Kocher, Hansjörg Baurecht, Benjamin Ehmke, Michael Nolde, Stefan Lars Reckelkamm

**Affiliations:** Institute of Health Services Research in Dentistry, University of Münster, Münster, Germany; Department of Restorative Dentistry, Periodontology, Endodontology, and Preventive and Pediatric Dentistry, University Medicine Greifswald, Greifswald, Germany; Department of Epidemiology and Preventive Medicine, University of Regensburg, Regensburg, Germany; Clinic for Periodontology and Conservative Dentistry, University of Münster, Münster, Germany

**Keywords:** Complement C3, immunomodulation, periodontitis, drug discovery, mendelian randomization analysis

## Abstract

**Aim:** Evidence from a phase IIa trial showed that a complement C3-targeted drug reduced gingival inflammation in patients with gingivitis. We investigated the therapeutic effect of genetically proxied C3 inhibition on periodontitis using drug target Mendelian randomization (MR).

**Materials and methods:** We used multiple ‘cis’ instruments from the vicinity of the encoding loci of C3 that are associated with serum C3. We selected three independent single nucleotide polymorphisms (rs141552034, rs145406915, rs11569479) from a genome-wide association study (GWAS) of 5,368 European descent individuals. We extracted association statistics from a GWAS of 17,353 clinical periodontitis cases and 28,210 European controls. Wald ratios were combined using inverse-variance weighted meta-analysis.

**Results:** MR analysis revealed that the inhibition of C3 reduces the risk of periodontitis (Odds ratio 0.91 per 1 standard deviation reduction in C3; 95% Confidence Interval 0.87–0.96, P-value=0.0003).

**Conclusions:** Findings from MR analysis showed that C3 blockade protects against periodontitis.

**Clinical relevance:** *Scientific rationale for study:* Mounting evidence suggests that the complement system is dysregulated in periodontitis. First human randomized clinical trials demonstrated promising effects mediated via the blockade of C3, a key factor of the complement system. Results from drug target MR analysis can provide compelling evidence to predict the efficacy of pharmacological C3 blockage in future clinical trials in periodontitis patients.

*Principle findings:* Genetically proxied C3 inhibition reduced periodontitis risk.

*Practical implications:* Our MR analysis provides genetic evidence that C3-targeted drugs might be an efficient adjunct therapy in periodontitis.

## Introduction

Periodontitis is a prevalent chronic inflammatory disease affecting around 50% of adults, of which about 10% suffer from a severe periodontitis (1). Dysregulated and excessive inflammatory response due to dysbiosis lead to the destruction of the supporting tissues around the tooth (e.g., gingiva, periodontal ligament, and alveolar bone) (2). When left untreated, periodontitis can lead to tooth loss, and, consequently, impaired mastication, and poor esthetics, which affects the patients’ quality of life (3). In addition to the direct impact on oral health, this condition is associated with an increased risk of other systemic conditions, including cardiovascular diseases, diabetes mellitus, rheumatoid arthritis, and Alzheimer’s disease (4). Thus, treating periodontitis may also reduce the risk of periodontitis-associated comorbidities. Periodontal therapy reduces infection and inflammation by mechanically removing dental plaque and calculus, often with adjunctive antimicrobials, while optimizing the patients’ biofilm control (5). However, in highly susceptible cases, the treatment is deemed ineffective (6). It has been hypothesized that host modulation therapy can be incorporated into the management of periodontitis (7).

The complement system is a critical part of the innate immune system. Recent studies have shown that it not only acts as a first-line defense mechanism against pathogens and endogenous molecules but also modulates the host immune response by engaging in crosstalk interactions (8). In addition, complement dysregulation or excessive activation (e.g., due to genetics or microbial virulence factors) becomes a pathological factor causing and exacerbating a number of disorders, including periodontitis. Observational human studies, which reported elevated complement metabolites in gingival fluid during periodontal tissue inflammation, support this presumed role in the pathogenesis of periodontitis (9). Moreover, treatment that alleviated periodontal inflammation was associated with reduced complement C3 activation in gingival crevicular fluid (10).

The complement cascade can be initiated by three distinct mechanisms: the classical, lectin, and alternative pathways. Inflammation caused by any of these pathways is mediated through C3 activation. Thus, C3 portrays a ‘functional hub’ where all complement activation pathways converge, rendering this protein a potential pharmacological target (see Figure 1) (11). Animal models revealed that C3-deficient mice did not develop gingival inflammation and alveolar bone loss due to periodontitis (12). Cp40, an analog to C3-inhibiting compstatin, blocks the binding of C3 to its convertase and thus interrupts the complement cascade. Cp40 treatment in mice and non-human primates led to a decrease in periodontal inflammation and tissue destruction (12–14). Consequently, Cp40 was clinically developed for human use as ‘AMY-101’ and revealed promising safety and efficacy endpoints in a phase I safety trial (11) and a phase IIa proof-of-concept study in gingivitis (15).

**Figure 1:**
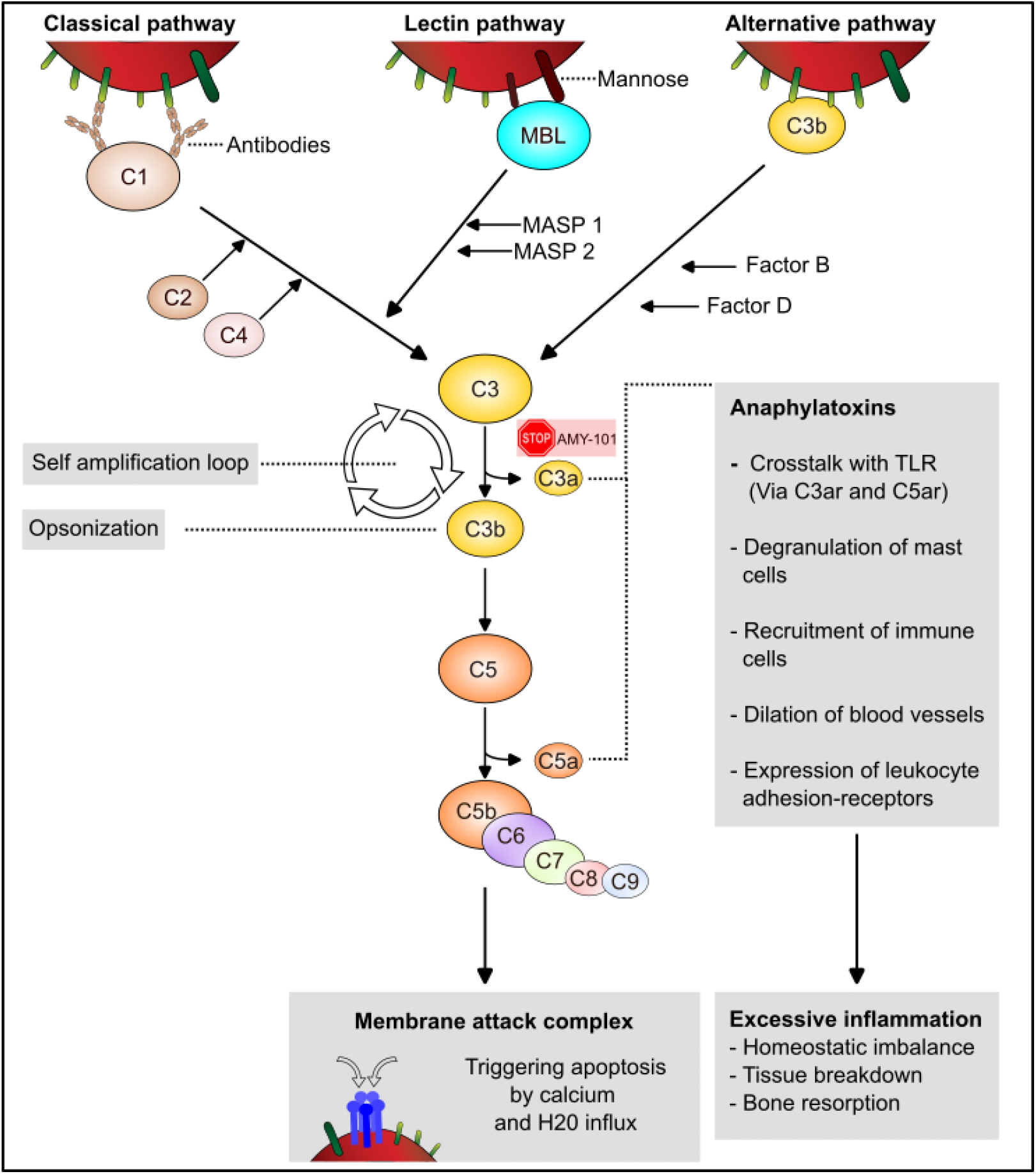
Simplified schematic of the complement cascade (inhibition) Three different pathways can activate the complement system. Classical activation occurs when antibodies bind to antigens and is mediated by complement factor C1. The lectin pathway requires the binding of mannose-binding lectin (MBL) to mannose residues of microbial polysaccharides. It is triggered in the absence of antibodies but follows the classical pathway. The alternative pathway begins with the spontaneous breakdown of C3 into C3a and C3b. C3b is rapidly inactivated unless a microbial surface is nearby. In this case, C3b and additional factors form the C3 convertase. Each of these pathways leads to the cleavage of the central complement C3, followed by the cleavage of C5 by so-called C5 convertases with the subsequent formation of a lytic membrane attack complex (MAC). The released C3a and C5a are highly potent anaphylatoxins and exert immunomodulatory and pro-inflammatory effects. Precisely these effects are associated with the excessive inflammation of the periodontium and the resulting pathophysiological changes (6).

In our study, we employed a Mendelian randomization (MR) approach to genetically test whether inhibiting C3 interferes in the pathogenesis of periodontitis. We used genetic variants that serve as instrumental variables for the druggable protein target ‘C3’ and studied the corresponding potential effects of AMY-101 in periodontitis. Despite preclinical and clinical studies showing potential safety and efficacy of AMY-101, the failure rate of candidate drugs progressing through clinical trials is high, with increasing failure rates in phase II and phase III (Holmes 2021). Our study thus aims to determine whether C3 is causative for periodontitis and may predict the outcome of future clinical trials in periodontitis patients.

## Materials and methods

MR is an instrumental variable approach to determine whether there is a causal relationship between a modifiable risk factor and a disease. The MR study design aims to draw causal inference minimizing the impact of bias from confounding and reverse causation that conventional observational studies suffer from. Genetic variants carried from parents to offspring are randomly allocated during conception; hence, independent of observed and unobserved confounders of the exposure-outcome association (16). Also, genetic associations are guarded against reverse causation since genetic variants remain fixed throughout one’s lifetime and are not altered by the disease process (17). This natural randomization makes MR analysis analogous to randomized clinical trials (see Figure 2). In genetic association studies, the measured effect corresponds to the presence of the ‘*effect allele’* in the genetic variant (equivalent to a drug in a randomized clinical trial), compared to a *‘baseline allele’* variant (similar to a placebo in a randomized clinical trial) (18). The standard MR approach selects single nucleotide polymorphisms (SNPs) from the whole genome regardless of the location of these variants with respect to the encoding loci. Drug target MR (or *cis*-MR) utilizes SNPs from within and around a gene known to encode a druggable protein, like C3. While standard MR elucidates the causal relationship between a modifiable risk factor and an outcome of interest, cis-MR determines whether altering a certain drug target changes the risk of a disease (19). To validly assess the causal relationship, instruments must satisfy the relevance, independence, and exclusion restriction/no-horizontal pleiotropy assumptions. The first assumption requires that the instruments are strongly associated with the risk factor. The second assumption states that there are no common causes of the instruments and outcome. The third assumption entails that the instruments alter the outcome only through the exposure (20).

**Figure 2:**
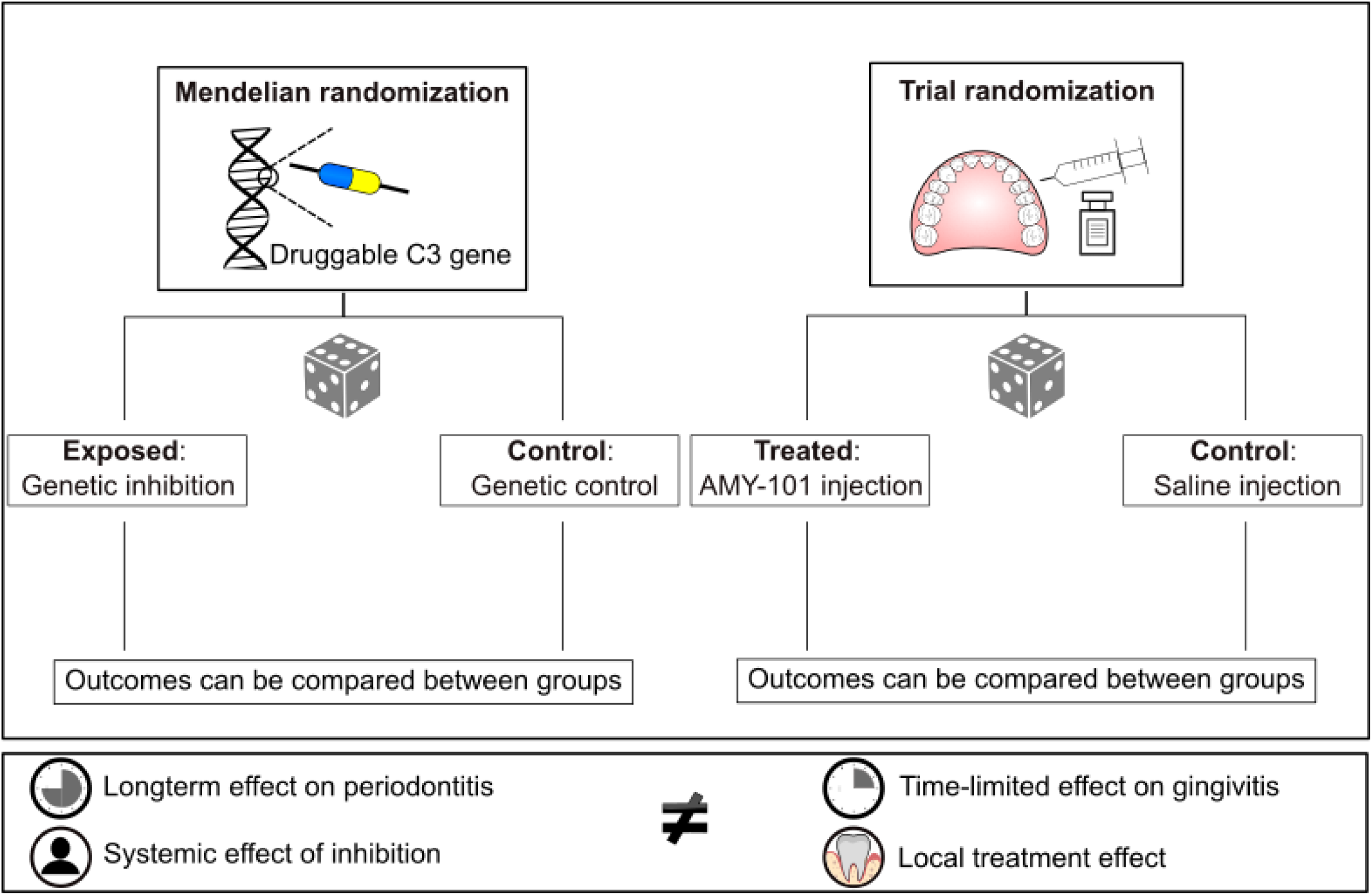
A comparison of study principles. This overview compares and contrasts parallels between the clinical trial of AMY-101 and our Mendelian Randomization (MR) approach. In the clinical trial, randomization was achieved by random allocation of drug-application sites, whereas in MR it was achieved by random allocation of alleles. The clinical trial examined the local and time-limited effect of AMY-101, while the MR approach examined the effect of lifelong exposure to a hypothetical drug (i.e. genetic variant) that targets the encoded protein (17).

### C3 indexing GWAS

Variants were derived from a genome-wide association study (GWAS) of serum concentration of C3 from a large-scale proteogenomic study (N=5,368) from the Age, Gene/Environment Susceptibility cohort. Association estimates in this GWAS were derived from participants of European descent. C3 in serum was detected and quantified by the Slow-Off rate Modified Aptamer proteomic profiling technology (21).

### Periodontitis GWAS

Summary statistics for periodontitis were obtained from the Gene-Lifestyle Interactions in Dental Endpoints consortium. A total of 17,353 participants of European ancestry were classified as clinical periodontitis cases and 28,210 as controls. Periodontitis was defined by either the Centers for Disease Control and Prevention / American Academy of Periodontology classification or the Community Periodontal Index case definition (22).

### Instrument selection

We identified cis-acting variants as proxies for C3 inhibition within a region 300 kB 5′ to 300 kB 3′ of the drug target encoding loci (chromosome 19; 6,677,715-6,730,573 (GRCh37/hg19)) that were associated with serum C3 levels (P-value < 1×10^−4^). We clumped the genetic variants by applying a conservative threshold (< 0.001) to assure that instruments are not in linkage disequilibrium. We estimated the F statistics to measure instrument strength. A magnitude of 10 is considered a reasonable threshold to rule out weak instrument bias (20). It should be noted that cis-acting variants have a substantial effect on protein expression compared to other potential downstream biomarkers and this ensures the validity of the relevance assumption. Furthermore, we searched the instruments in Phenoscanner to confirm their independence from any known confounders (23). Because it is a protein-disease relationship rather than a downstream biomarker-disease relationship, our MR study is less prone to violate the assumption of ‘no horizontal pleiotropy’ (19).

### Statistical analysis

Data for the exposure and outcome were harmonized according to the effect allele and none of the instruments were palindromic. The individual causal effect for each instrument was calculated as the ratio of the instrument-outcome association to the instrument-exposure association. Ratio estimates were pooled using inverse variance weighted (IVW) meta-analysis. Additionally, we performed a leave-one-out analysis to assess if the causal effect substantially changes upon removal of a single instrument (20). All analyses were performed in R version 4.1.2 using the TwoSampleMR and MendelianRandomization packages.

#### Ethics

All analyses were based on publicly available summary statistics without accessing individual level data; hence, ethical approval was not required. The included GWAS received informed consent from the study participants and were approved by pertinent local ethical review boards.

## Results

Three single nucleotide polymorphisms (rs11569479, rs141552034, rs145406915) were employed as instrumental variables to convey the effect of genetically proxied C3 blockade on periodontitis risk (Supplementary Table 1). F-statistics ranged between 13-45, indicating no weak instrument bias. None of the SNPs were associated with any common cause of C3 and periodontitis. Our MR analysis found that the inhibition of C3 reduces periodontitis risk (Odds ratio 0.91 per 1 standard deviation reduction in C3; 95% Confidence Interval (CI) 0.87–0.96) (Table 1). In a leave-one-out analysis, we showed that the direction of effect was not primarily influenced by a single instrument (Supplementary figure 1).

**Table 1.** Mendelian Randomization Estimate for effects of complement C3 Inhibition on outcomes using genetic variants as Instruments

| Outcome | Method | No. SNPs | OR | CI | P Value |
| --- | --- | --- | --- | --- | --- |
| Periodontitis risk | IVW | 3 | 0.91 | 0.865;0.958 | 0.0003 |

OR, odds ratio per one standard deviation increase in C3. CI, Confidence interval. IVW, inverse-variance weighted analysis based on genetic associations of 3 uncorrelated SNPs (rs11569479, rs141552034, and rs145406915) with complement C3 and periodontitis risk

## Discussion

The present study used a *cis*-MR approach to investigate the potential therapeutic effect of C3 blockade on periodontitis. Our results showed that downregulation of C3 lowered periodontitis risk. This finding is in line with proof-of-concept studies of C3 inhibition in preclinical models of periodontitis (12–14) and with a randomized controlled clinical trial (phase IIa) of C3 inhibition in patients with gingivitis (15).

The first in-human clinical trial recruited 50 healthy males to investigate the safety and tolerability of a single ascending dose and multiple doses of AMY-101 (11). Up to 21 days after treatment, none of the participants experienced treatment-related adverse events. Also, the pharmacokinetic and pharmacodynamic profile of AMY-101 proved its suitability for further testing in clinical trials (11). In 2019, the US Food and Drug Administration approved AMY-101 as an investigational new drug, for a Phase IIa clinical trial to evaluate its safety and efficacy in patients with gingival inflammation (15). 40 participants (50% female) were included in a placebo-controlled, double-blinded, split-mouth study design. In the dose selection phase, 12 participants were randomized to three dosing groups (0.025, 0.1, and 0.5 mg per interdental papilla) to identify a safe and effective dose. Consequently, 0.1 mg per interdental papilla was chosen for the main study and given to the efficacy population of 32 patients. All participants (N=40), regardless of the AMY-101 dose, represented the safety population and demonstrated the desired safety profile. The inflammation-related clinical indices, ‘modified gingival index’ (MGI) and ‘bleeding on probing’ (BOP), served as primary and secondary outcomes, respectively. Four weeks after treatment initiation, the MGI (measured at six sites on the tooth) showed a greater improvement in the treatment group than in the placebo group (least squares mean difference of – 0.181, 95% CI: –0.248 to –0.114). Similarly, BOP was greatly reduced in the treated sites (15). Tissue destruction-related clinical measures, such as pocket depth (PD) or clinical attachment loss (CAL), form the basis of periodontitis diagnosis (24) and did not change greatly from baseline measurements. This observation can be due to the lack of the minimum number of participants needed to see the clinically relevant changes. In our study, the outcome of interest was periodontitis risk; thus, it demonstrates the impact of C3 inhibition on a broader range of clinical indices including PD and CAL. In comparison to our result, preclinical studies revealed less alveolar bone loss in sites injected with AMY-101 than in placebo-treated sites (12). These preclinical features of C3 blockade, in addition to our findings, warrant further investigation of AMY-101 in clinical trials.

The complement system is an important and powerful actor in the host defence system. However, recent discoveries showed that its excessive stimulation causes tissue damage. Once activated, the complement system follows a cascade of opsonisation of the target (e.g. a bacterial cell), self-amplification, generation of effector molecules and immune crosstalk (see Figure 1) (25). C3, a central node in all relevant pathways, presents itself as a promising therapeutic target (see Figure 1). In 2007, the first C3 inhibitor ‘pegcetacoplan’ was approved for use in patients with nocturnal hemoglobinuria. This discovery revealed the relevance of this treatment strategy and set the scene for the development of therapeutic C3 antagonists in other diseases like retinal diseases, neurodegenerative diseases, severe coronavirus disease, and periodontitis (26). AMY-101 still has to overcome phase IIb (to set the optimal dose to show biological activity with minimal side effects) and phase III (to assess the therapeutic effectiveness) before being approved for patient use. Acknowledging that the main issue in drug development is failure due to lack of efficacy in phases II and III (27), findings from MR studies provide compelling evidence of the causal relationship between protein drug targets and diseases, increasing the probability of candidate drugs to succeed in phase III clinical trials (28). On several occasions MR studies predicted therapeutic effects of candidate drugs prior to their testing in clinical trials (27). In the phase IIa clinical trial by Hasturk et al., randomization was achieved by the random allocation of drug-application sites, whereas in our MR the random allocation of the genetic variants acts as a randomization device. A key difference, however, is that whereas the phase IIa clinical trial examined the local short-term effect of AMY-101, this drug-target MR study examined the long-term inhibition of C3 (17).

A key strength of our study is that we applied a protein drug target MR analysis. In an attempt to ensure the validity of our instruments and minimize the risk of violating the no horizontal pleiotropy assumption, instrumental variables were identified based on their proximity to the C3 gene and in relation to C3 levels rather than an association with downstream biomarkers. Nevertheless, the study has limitations. First, the exposure and outcome association estimates were derived from individuals with European ancestry. Linkage disequilibrium patterns can differ between populations and may not extend to other ethnic groups, therefore limiting the generalizability of our findings to other ethnicities (20). Although plausibly selected, we could not validate our instrumental variables based on mRNA expression due to the unavailability of such data. Finally, since we selected the SNPs from the vicinity of a single gene region, we were unable to apply pleiotropy-robust MR methods.

## Conclusion

Drug target MR facilitates an early assessment of a drug candidate and is gaining interest as a fundamental tool in drug development. Our study suggests a beneficial effect of pharmacologically targeting C3 in periodontitis and recommends further testing of AMY-101 in a phase III clinical trial.

## Data Availability

Summary genetic data for complement C3 protein are deposited at the GWAS Catalog with accession ID: GCST90088016. The periodontitis summary data are available at https://data.bris.ac.uk/data/dataset/2j2rqgzedxlq02oqbb4vmycnc2

https://www.ebi.ac.uk/gwas/studies/GCST90088016

## Data availability

Summary genetic data for complement C3 protein are deposited at the GWAS Catalog with accession ID: *GCST90088016*. The periodontitis summary data are available at https://data.bris.ac.uk/data/dataset/2j2rqgzedxlq02oqbb4vmycnc2

