## Supplementary table and figure for "Using human genetics to validate complement C3 as a drug target in periodontitis": 20230208_C3_Perio_Supp.docx

|  |  |  |  | Estimates for C3 | | |  | Estimates for Periodontitis | | |
| --- | --- | --- | --- | --- | --- | --- | --- | --- | --- | --- |
| SNP | EA | OA | EAF | Beta | SE | P-value | F | Beta | SE | P-value |
| rs11569479 | T | C | 0.868 | 0.183 | 0.028 | 9.5e-11 | 42.1 | 0.016 | 0.023 | 0.475 |
| rs141552034 | G | T | 0.972 | 0.218 | 0.057 | 1.3e-04 | 14.7 | 0.042 | 0.086 | 0.626 |
| rs145406915 | A | C | 0.976 | 0.231 | 0.063 | 2.4e-04 | 13.5 | -0.009 | 0.159 | 0.955 |

Supplementary Table 1. Associations of genome-wide significant single nucleotide polymorphisms (SNPs) for Complementary Component C3 from the GWAS by Gudjonsson et al. (Gudjonsson et al. 2022) and periodontitis from the GWAS by Shungin et al. (Shungin et al. 2019)

EA, effect allele. OA, other allele. EAF, effect allele frequency. Beta, regression coefficient. SE, standard error. F, F statistics

Supplementary Figure 1: Leave-one-out IVW Analyses

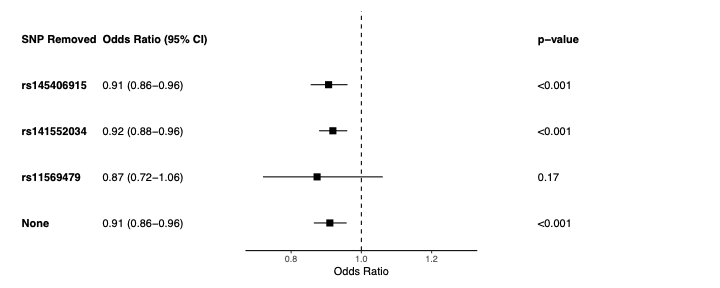
